# Imported malaria predominates in near-elimination settings in Southwestern Uganda

**DOI:** 10.64898/2026.01.25.26344810

**Authors:** Monica Mbabazi, Shahiid Kiyaga, Thomas Katairo, Kisakye D. Kabbale, Victor Asua, Brian A. Kagurusi, Innocent Wiringilimaana, Bienvenu Nsengimaana, Francis D. Semakuba, Jackie Nakasaanya, Alisen Ayitewala, Eric Watyekele, Isaiah Nabende, Tonny Max Kayondo, Emmanuel Arinaitwe, Jerry Mulondo, Stephen Tukwasibwe, Samuel L. Nsobya, Bosco Agaba, Catherine Maiteki, Daudi Jjingo, David Patrick Kateete, Moses R. Kamya, Isaac Ssewanyana, Andres Aranda-Diaz, Melissa D. Conrad, Maxwell Murphy, Inna Gerlovina, Adrienne Epstein, Isabel Rodriguez-Barraquer, Philip J. Rosenthal, Grant Dorsey, Bryan Greenhouse, Jessica Briggs

## Abstract

**Background:** Malaria transmission in southwestern Uganda is low, but persists despite ongoing control efforts. Identifying whether infections are locally sustained or imported by travelers is critical for guiding interventions. We integrated epidemiologic surveillance with parasite genomics to characterize imported malaria episodes at three health facilities in southwestern Uganda.

**Methods:** Between January 2023 and June 2024, we enrolled microscopy-confirmed malaria cases at three health facilities, Maziba and Muko (very low transmission) and Kamwezi (low-to-moderate transmission), administered travel history questionnaires, and collected dried blood spots for genotyping. *Plasmodium falciparum* infections were genotyped using MAD^4^HatTeR, a highly sensitive multiplex amplicon sequencing panel targeting 165 diversity markers and 38 drug resistance loci. Complexity of infection and pairwise relatedness were estimated using MOIRE and Dcifer, respectively. Plasmotrack, a Bayesian transmission network framework, was used to infer network structure, transmission directionality, reproduction numbers, and importation rates.

**Results:** Amongst malaria cases, recent overnight travel was common in Maziba (87%) and Muko (96%) but infrequent in Kamwezi (12%). Most travel in cases from Maziba and Muko was from high-transmission regions in northern and eastern Uganda. Parasites in Maziba and Muko cases exhibited higher within-host diversity and lower within-site relatedness compared to those in Kamwezi cases. Transmission network inference identified most infections in Maziba and Muko as imported, with the majority of inferred secondary transmission linked to recent travelers. In contrast, Kamwezi showed multiple highly related clusters, indicating sustained local transmission. Validated and candidate markers of artemisinin partial resistance (K13 P441L and R561H) were more prevalent in Kamwezi.

**Conclusion:** Malaria in Maziba and Muko was driven largely by importation from other parts of Uganda, while local transmission played a larger role in Kamwezi . Tailored interventions addressing travel-associated risks and local transmission, supported by travel histories and parasite genetic data will be valuable to advance malaria elimination in this region.

## Introduction

Malaria, caused principally by *Plasmodium falciparum,* remains a major public health challenge across sub-Saharan Africa, despite decades of sustained control efforts [1]. In Uganda, which ranks third in global malaria case burden, most regions experience moderate to high transmission intensity. However, the southwestern part of the country, particularly highland areas, has historically experienced much lower transmission. According to the Uganda Ministry of Health 2018-2019 Malaria Indicator Survey [2], parasite prevalence by microscopy in children < 5 years of age was below 5% in several southwestern districts, compared to over 30% in northern and eastern regions. This unique epidemiological profile has made southwestern Uganda a prime candidate for subnational elimination, and the Uganda National Malaria Elimination Division (NMED) has prioritized it for targeted interventions. However, the optimal interventions to achieve elimination remain unclear because the factors sustaining residual transmission in this setting are unknown.

Low malaria transmission regions face distinct challenges compared to high-burden regions. Transmission in these settings tends to be focal and heterogeneous, requiring interventions to be appropriately targeted to be efficient and effective [3]. Two factors particularly complicate malaria control and elimination efforts in low-transmission contexts: epidemic potential and human mobility. Highland areas with cooler temperatures, such as those found in southwestern Uganda, have historically experienced low or unstable malaria transmission, leaving populations with limited acquired immunity. This renders these populations vulnerable to epidemic transmission and the potential for frequent severe disease when conditions permit, making these regions particularly challenging for malaria control [4,5]. Weather patterns, such as extreme precipitation and flooding, and environmental changes, including land use modifications, deforestation, and climate warming, can increase malaria transmission in highland areas, contributing to epidemic potential [4,6–9]. Another well-documented challenge in low-transmission settings, is the role of human mobility, potentially leading to imported infections. Studies across Africa and Southeast Asia have shown that even sporadic importation can sustain or reignite transmission, overwhelm surveillance systems, and obscure the distinction between local and imported cases [10–12]. Critically, importation is not limited to international travel; within-country movement between regions of differing transmission intensity can reintroduce malaria parasites into areas with otherwise minimal transmission [13,14]. As evidence of this, in a prior cohort study conducted across three sites with differing transmission intensities in Uganda, recent overnight travel increased malaria risk 3.5-fold, with blood smear positivity rising from 2.2% before travel to 7.8% within 30 days of return [15].

These dynamics are particularly relevant in southwestern Uganda, a low transmission highland region that borders Rwanda, where transmission is generally moderate, and the Democratic Republic of Congo, which bears a high and sustained malaria burden [16–18]. Frequent human movement across borders and/or within Uganda may render southwestern Uganda particularly vulnerable to importation-driven transmission. For example, a case-control study of symptomatic highland residents in southwestern Uganda showed that those who had recently traveled to a higher transmission area were nearly seven times more likely to be diagnosed with malaria than those who had not [13]. Introductions of malaria by travelers have the potential to sustain transmission, particularly in areas where surveillance systems may miss low-level or asymptomatic infections [19]. In addition, if malaria in these settings is mostly acquired from other regions, intensification of local vector control may provide limited benefit. Understanding how imported infections may influence malaria transmission is critical to tailoring effective control and elimination strategies.

In recent years, advances in generating and analyzing genomic data have offered powerful tools to investigate malaria transmission dynamics with greater resolution [20–22]. By analyzing parasite genetic relatedness, it is increasingly possible to distinguish imported from local infections [23–25]. In this study, we integrated sensitive, diverse multiplexed amplicon sequencing with patient demographic and travel history data to investigate *P. falciparum* transmission at three health facilities in southwestern Uganda. Our aim was to better understand the contribution of imported infections to malaria transmission in Southwestern Uganda and to inform the design of control interventions appropriate for settings with low-level malaria transmission.

## Materials and Methods

### Ethical statement

This study was approved by the Makerere University School of Biomedical Sciences Research and Ethics Committee (SBS-2021-163), the Uganda National Council for Science and Technology (HS2309ES), and the University of California, San Francisco Institutional Review Board (Ref No. 341609). Written informed consent was obtained from all adult participants (aged 18 years and above). In accordance with Uganda’s National Guidelines for Research Involving Humans as Research Participants, assent was obtained from all children aged 8 years and older, and written consent was obtained from their parents or guardians prior to enrollment. For children below 8 years of age, only parental or guardian consent was obtained.

### Study design and data collection

This study was conducted within the Implementing Malaria MoleculaR SurveillancE in Uganda (IMMRSE-U) project at three health facilities in southwestern Uganda: Maziba Health Center IV (Kabale District), Muko Health Center IV (Rubanda District), and Kamwezi Health Center IV (Rukiga District) (**Fig 1**). In addition to primary data collection, we accessed enhanced malaria surveillance data from the Uganda Malaria Surveillance Program (UMSP) [26], including individual-level case data with demographic information and site-level incidence and test positivity rate (TPR) metrics over the study period. These data provided contextual information on malaria burden and trends at the study health facilities, enabling comparison of study participants with the broader patient population attending the health facility and supporting interpretation of site-specific transmission patterns.

**Fig 1.**
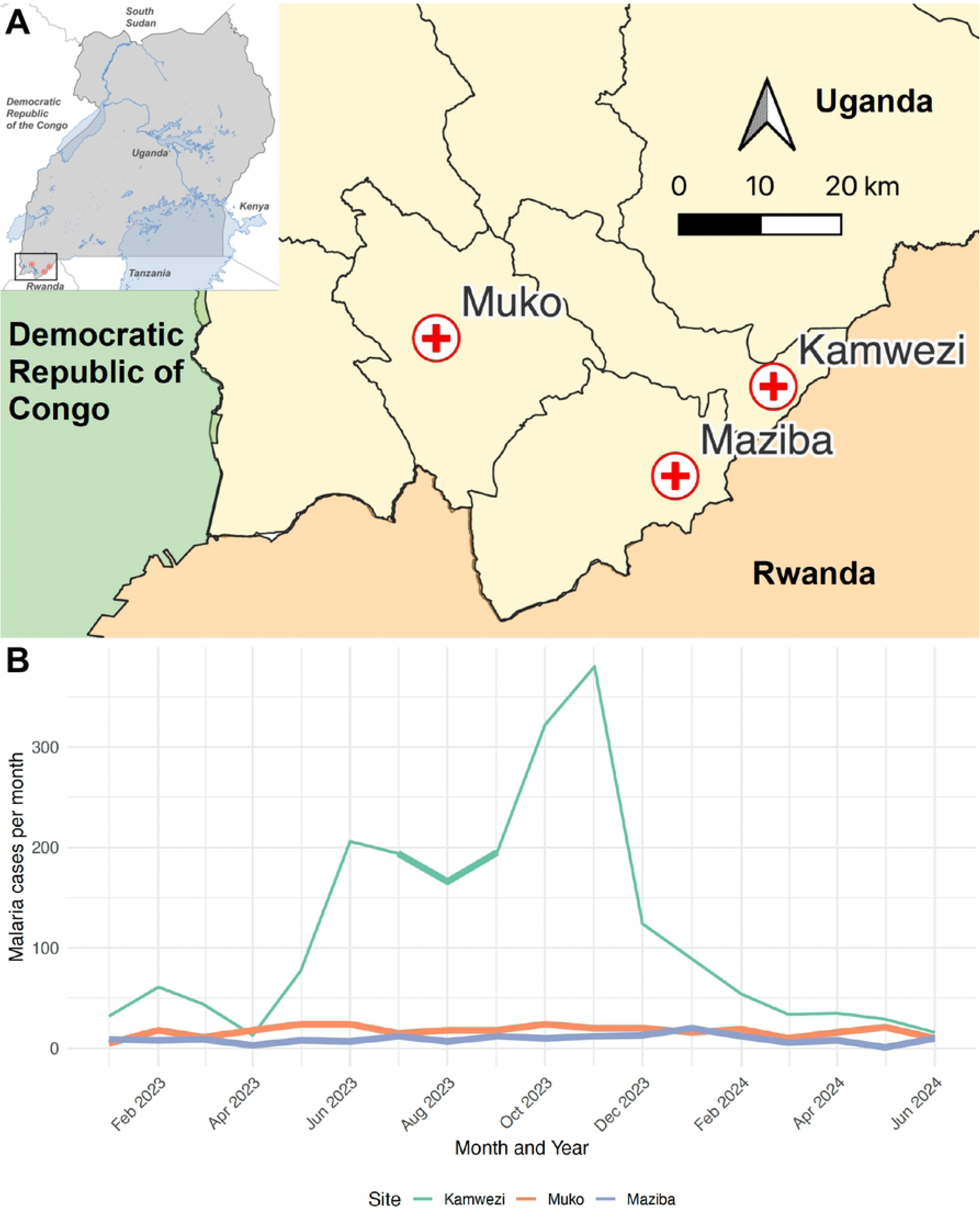
Study sites and monthly confirmed malaria cases at each site over the course of the study. (A) Map of Uganda showing the location of the study region (inset, top left). The inset highlights the area containing the study sites, which is shown at higher resolution in the main map panel. The main panel displays the three study health facilities, Maziba Health Center IV, Muko Health Center IV, and Kamwezi Health Center IV, in southwestern Uganda. Distances on the map are shown in kilometers. (B) Monthly confirmed malaria cases recorded at each study health facility according to the Uganda Malaria Surveillance Program (UMSP). The time period of sample collection is highlighted with thicker lines in B, showing a 2-month period of sample collection at Kamwezi and an 18-month period of sample collection at Maziba and Muko.

In the very low transmission settings of Maziba and Muko, we aimed to enroll and genotype all microscopy-confirmed malaria cases throughout the study period from 1^st^ January 2023 to 30^th^ June 2024. Consented participants completed a standardized travel history questionnaire and provided a dried blood spot (DBS) sample for parasite genomic analysis. During the study period, routine surveillance data from the Uganda Malaria Surveillance Program (UMSP) recorded a total of 474 microscopy-confirmed malaria cases at Muko (n = 307) and Maziba (n = 167). Although we aimed to enroll all cases; dried blood spot (DBS) samples were ultimately available for 56.0% (172/307) of cases in Muko and 56.8% (95/167) of cases in Maziba; these samples were subsequently processed for genomic analysis. Some enrolled cases also had missing travel-history data, and a small number of individuals completed travel questionnaires without a corresponding DBS sample collected, resulting in minor discrepancies between the totals available for epidemiologic versus genomic analyses. In Kamwezi, a setting of low-to-moderate transmission, 200 patients with microscopy-confirmed malaria were enrolled between July and August 2023. All participants enrolled at Kamwezi completed travel history questionnaires and provided DBS samples. Of these, 100 samples were randomly selected for genotyping. All DBS samples from the three sites were transported to the Central Public Health Laboratory (CPHL) in Butabika, Uganda, for processing and sequencing. The travel questionnaire captured demographic and clinical data, including age, sex, village and district of residence, occupation, and date of interview. Participants were also asked to report on overnight travel within the 30 days preceding their malaria diagnosis. This included whether they had traveled overnight outside their village, the destination(s) of travel (within or outside their home district, as well as outside Uganda), the duration of travel (departure and return dates), the primary reason for travel (e.g., work, visiting relatives, other), and the frequency of travel. These data were used to characterize patient movement patterns and explore their association with malaria transmission dynamics at each site.

## Laboratory Methods

### Sample processing, DNA extraction, and quantitative PCR

At CPHL, the DBS were accessioned into a sample database. DNA was extracted from DBS using the Tween-20/Chelex 100 method [27]. Extracted DNA was stored at -20 °C for subsequent analysis. To estimate *Plasmodium falciparum* parasite density, we utilized an ultrasensitive quantitative PCR (qPCR) assay targeting the *varATS* gene [28]. Parasite densities were determined by comparison with a standard curve of cultured ring-stage parasites diluted in whole blood at known parasitemia levels ranging from 1 to 10,000 parasites/microliter. Assay quality control was ensured by requiring an R² > 0.95 for control CT values on each plate.

### Library preparation and sequencing

Amplicon libraries were prepared using the Multiplex Amplicons for Drug, Diagnostic, Diversity, and Differentiation Haplotypes using Targeted Resequencing (MAD⁴HatTeR) assay, a highly sensitive multiplexed sequencing protocol designed to amplify 165 microhaplotypes for parasite diversity and 118 key markers in 38 genes for drug and diagnostic resistance surveillance [29]. DNA from dried blood spots were amplified using the MAD⁴HatTeR protocol as previously described (Aranda-Díaz et al., 2025) using primer pools D1.1/R1.2 and R2.1 [30]. Amplified libraries were purified with CleanMag Beads, indexed, and pooled by parasitemia strata to balance sequencing depth across libraries. Final pools were quality-checked using an Agilent Bioanalyzer and further bead-purified to remove residual primer dimers. Sequencing was performed on an Illumina MiSeq platform, generating 150 bp paired-end reads with a 5% PhiX spike-in.

### Bioinformatics analysis and quality control

Raw sequencing data were processed using the MAD⁴HatTeR amplicon sequencing pipeline, a publicly available Nextflow-based bioinformatics pipeline to filter, demultiplex, and infer alleles from FASTQ files, as previously described [29,31]. Data was processed with release 21 (v0.2.1) of the pipeline. Downstream analysis was performed using R (version 4.1.2). Data were merged across sequencing runs and sample- and allele-level quality filters were applied as follows. Samples in which > 50% of amplicons had < 100 reads were excluded, and alleles with within-sample allele frequency (WSAF) < 1% were excluded. Following these filters, 348 of 367 samples (94.8%) were retained for downstream analyses. Maziba contributed 95 genotyped samples, of which 89 passed QC (93.7%); Muko contributed 172 samples, of which 159 passed QC (92.4%); and all 100 samples from Kamwezi passed QC (100%) (**Table 1**). Median parasite density was higher among samples passing QC compared to those failing QC (11,216 parasites/µL vs. 3,978 parasites/µL, respectively).

**Table 1.** Demographic and travel behaviour of malaria cases across three study sites representing low and moderate transmission settings in southwestern Uganda.

| Dataset | Variable | Maziba | Muko | Kamwezi |
| --- | --- | --- | --- | --- |
| **Health Facility**<br>**(UMSP)** | No. of malaria cases | 167 | 307 | 2071 |
|  | Mean age | 25.3 | 26.6 | 18.6 |
|  | Male % | 50.9 | 65.1 | 43.9 |
|  | Malaria incidence per 1000 PY | 8.8 | 10.2 | 90.9 |
|  | Test Positivity Rate (TPR) | 12.7 | 14.0 | 31.7 |
| **Travel Survey** | No. of samples with travel survey data | 103 | 164 | 1138 |
|  | Reported overnight travel in last 30 days (%) | 87.4 | 96.3 | 12.0 |
|  | Reported overnight travel **outside** district (%) | 85.4 | 91.5 | 6.9 |
|  | Reported overnight travel outside village but **within** district (%) | 1.9 | 4.9 | 3.6 |
|  | International % | 0 | 0 | 1.5 |
|  | Primary reasons for travel: |  |  |  |
|  | Work/trade | 44 (49.4%) | 103 (66%) | 13 (9.7%) |
|  | Visiting relatives | 26 (29.3%) | 27 (17.3%) | 77 (57.5%) |
|  | Burial rites | 7 (7.9%) | 12 (7.7%) | 14 (10.4%) |
| **Genomic Data** | No. of samples genotyped | 95 | 172 | 100 |
|  | Genotyped samples that passed QC (%) | 89 (93.7%) | 159 (92.4%) | 100 (100%) |
|  | Genotyped samples with travel survey data | 68 (76.4%) | 104 (65.4%) | 100 (100%) |

## Data analysis

### Drug resistance polymorphisms

Single nucleotide polymorphisms (SNPs) known to be associated with drug resistance were output directly from the MAD⁴HatTeR bioinformatic pipeline. SNPs were called when allele coverage was ≥ 10 reads, and WSAF was ≥ 1.0%. The drug-resistance targets assessed included loci in the chloroquine resistance transporter (*Pf*CRT), dihydrofolate reductase (*Pf*DHFR), dihydropteroate synthase (*Pf*DHPS), multidrug resistance protein 1 (*Pf*MDR1), and the kelch13 propeller domain (*Pf*K13) protein. Mutation prevalence was calculated by classifying both pure mutant and mixed genotypes as mutants. To assess differences in the prevalence of *Pf*K13 mutations (P441L and R561H) across study sites, we constructed contingency tables using estimated mutant and wild-type counts and performed Fisher’s exact tests. Overall site differences were tested, and pairwise comparisons were performed using 2×2 Fisher’s tests with a significance threshold of p < 0.05.

### Parasite genetic diversity, relatedness, and network analysis

We quantified within-host parasite diversity and allele frequencies (including expected heterozygosity) using MOIRE, an R package implementing a Bayesian framework for polyallelic data [32,33]. MOIRE jointly estimates allele frequencies, complexity of infection (COI, the number of genetically distinct parasite strains infecting an individual), and within-host relatedness while explicitly accounting for genotyping error. MOIRE also estimates effective complexity of infection (eCOI), a continuous measure that integrates COI and within-host relatedness.

Dcifer v1.2,1, an identity-by-descent (IBD)-based method designed to estimate genetic distance between polyclonal infections, was used to calculate pairwise relatedness (*r*) between infections [34,35]. Dcifer explicitly incorporates COI and population allele frequencies estimated from MOIRE, as well as unphased multiallelic data such as microhaplotypes, providing a statistically principled estimate of pairwise relatedness. The relatedness parameter r represents the sum of identity-by-descent shared across all parasite strains present in the two infections, capturing the total proportion of the genome inherited from recent common ancestors [35]. Pairwise relatedness of infections was computed with the *ibdDat()* function implemented in Dcifer. Statistical significance of relatedness was assessed using a one-sided hypothesis test with a null relatedness parameter of *r* > 0.125, representing approximately 3 generations of full outcrossing from a common ancestral parasite. Resulting p-values were corrected for multiple comparisons using the Benjamini–Hochberg (BH) false discovery rate procedure. Highly related pairs were defined as pairs with BH-adjusted p-values < 0.05. We then quantified: (i) the proportion of highly related pairs within each site, and (ii) the distribution of IBD values (mean, median) across all pairwise comparisons.

Networks of related infections were reconstructed using two complementary approaches. First, highly related infection pairs identified with Dcifer were used to generate undirected cluster networks in which each sample was represented as a node and edges connected highly related pairs, as defined above. As an additional approach, we applied a novel probabilistic transmission network model (Plasmotrack) to the study data [36]. Plasmotrack uses a Bayesian framework to infer transmission networks from genomic and epidemiological data, enabling estimation of key epidemiologic parameters including effective reproduction numbers, importation rates, and transmission directionality [36]. For both Dcifer and Plasmotrack networks, we integrated genomic data with survey data to characterize the epidemiologic characteristics of identified clusters.

Using the output from Plasmotrack founder infections (infections with no observed parents) and source infections (founders with mean effective reproduction number R_e_ > 0.8) were identified. Associations between inferred transmission network roles (i.e., founder or source infections) and epidemiologic traits were assessed using statistical tests. Fisher’s exact test was used to evaluate whether founder infections were more prevalent among individuals reporting recent travel compared to those reporting no travel.

### Statistical analysis

To assess differences in parasite complexity of infection (COI and eCOI) across study sites, we first performed pairwise Wilcoxon rank-sum tests. P-values were reported both with and without adjustment for multiple comparisons using the BH method. We used linear regression models to test for associations between infection complexity (COI and eCOI) and individual travel history. Travel was categorized as “no travel,” “inside district,” or “outside district” based on survey responses. To investigate whether the transmission intensity at reported travel destinations was associated with infection complexity, we linked participants’ travel destinations to Malaria Atlas Project (MAP) modeled annual parasite incidence data [37]. Both univariate and multivariate regression models were fitted to assess associations between destination incidence (low vs. moderate-high) and infection complexity, with additional models including site as a covariate.

## Results

### Participant demographics and travel history by site

Malaria burden varied substantially across the three study sites, with incidence and TPR from the health facilities indicating substantially lower malaria transmission in Maziba and Muko compared to Kamwezi (**Table 1**). Demographic characteristics also varied by site; individuals presenting with malaria were older and more likely to be male in the low transmission sites (Maziba and Muko) compared to Kamwezi.

Marked differences were observed in reported travel behaviors. Overnight travel in the last 30 days was reported by nearly all participants in Maziba and Muko (87.4% and 96.3%, respectively), but by only a small minority in Kamwezi (12.0%). The majority of trips in both Maziba and Muko were to destinations outside the district. Although far fewer participants in Kamwezi reported travel, those who did also predominantly traveled outside the district (6.9% outside versus 3.6% within the district) (**Table 1**). A history of cross-national travel was uncommon, and reported only in Kamwezi at a very low rate (1.5%). The primary reason for travel also differed by site: work and trade dominated in Maziba and Muko, while visiting relatives was the leading reason in Kamwezi. Among participants reporting travel, destinations extended beyond southwestern Uganda to northern and eastern regions of the country, which experience substantially higher malaria transmission (**Fig S1**). Participants from Maziba and Muko traveled substantially farther than those in Kamwezi, with median (Q1-Q3) travel distances of 273 km (203–345) and 221 km (130–314), respectively, compared with 70.5 km (16–136) in Kamwezi. These quantitative estimates are consistent with the long-distance mobility patterns illustrated in the mapped travel destinations (**Fig S1**).

### Parasite genetic diversity and relatedness

Genomic analyses revealed distinct patterns of parasite diversity and relatedness across the three study sites **(Table 2)**. The mean complexity of infection (COI) was highest in Muko (2.4), followed by Maziba (2.1), and Kamwezi (1.7). Similar trends were observed for effective complexity of infection (eCOI) (**Fig S2**) and the proportion of polyclonal infections (**Table 2**). We also observed greater heterogeneity in the distributions of COI and eCOI in Maziba and Muko compared to Kamwezi. Regression analyses found no significant associations between individual-level COI and either recent overnight travel or malaria transmission intensity at travel destinations in univariate models or in multivariate models adjusting for site.

**Table 2.** Summary of parasite genetic diversity and relatedness metrics across study sites.

| Variable | Maziba | Muko | Kamwezi |
| --- | --- | --- | --- |
| No. of samples | 89 | 159 | 100 |
| Mean COI | 2.1 | 2.4 | 1.7 |
| Mean effective COI | 1.7 | 1.9 | 1.4 |
| Polyclonal infections (%) | 51.7 | 65.5 | 44.0 |
| Pairs of highly-related samples (%) | 0.59 | 0.17 | 6.36 |
| Mean $H_e$ (top 20 loci) | 0.81 | 0.83 | 0.79 |
| Mean pairwise IBD | 0.015 | 0.014 | 0.065 |

In contrast, between-host relatedness, measured as mean pairwise identity-by-descent (IBD), was substantially higher in Kamwezi compared to Maziba and Muko (**Table 2**). The proportion of highly related infection pairs also followed this pattern, with Kamwezi showing the greatest number of highly related infections, whereas Maziba and Muko had very few highly related pairs. Estimated heterozygosity (H_e_) was similar across the three sites, with mean H_e_ values for the top 20 most diverse markers ranging from 0.79 to 0.83 across sites, indicating comparable levels of allelic diversity at the parasite population level.

### Clustering and Transmission Network Analysis

Analysis of pairwise identity-by-descent (IBD) revealed distinct clusters of highly related infections at each site (defined as r > 0.125, **Figure S3**). There were very few infections that were highly related between sites (3 highly related infection pairs between Maziba and Kamwezi, and 1 highly related infection pair between Maziba and Muko). To investigate within-site clusters, we created networks based on IBD (with undirected edges) and by using a transmission network model developed for *P. falciparum* (with directed edges, **Fig 2**). In Maziba, five small clusters were identified, and approximately 7% of samples belonged to at least one cluster. The largest cluster contained 6 related infections. In Muko, ten small clusters were detected (each ≤ 4 infections), and approximately 3% of samples were part of at least one cluster. Therefore, in Maziba and Muko, most infections were unrelated to each other.

**Fig 2.**
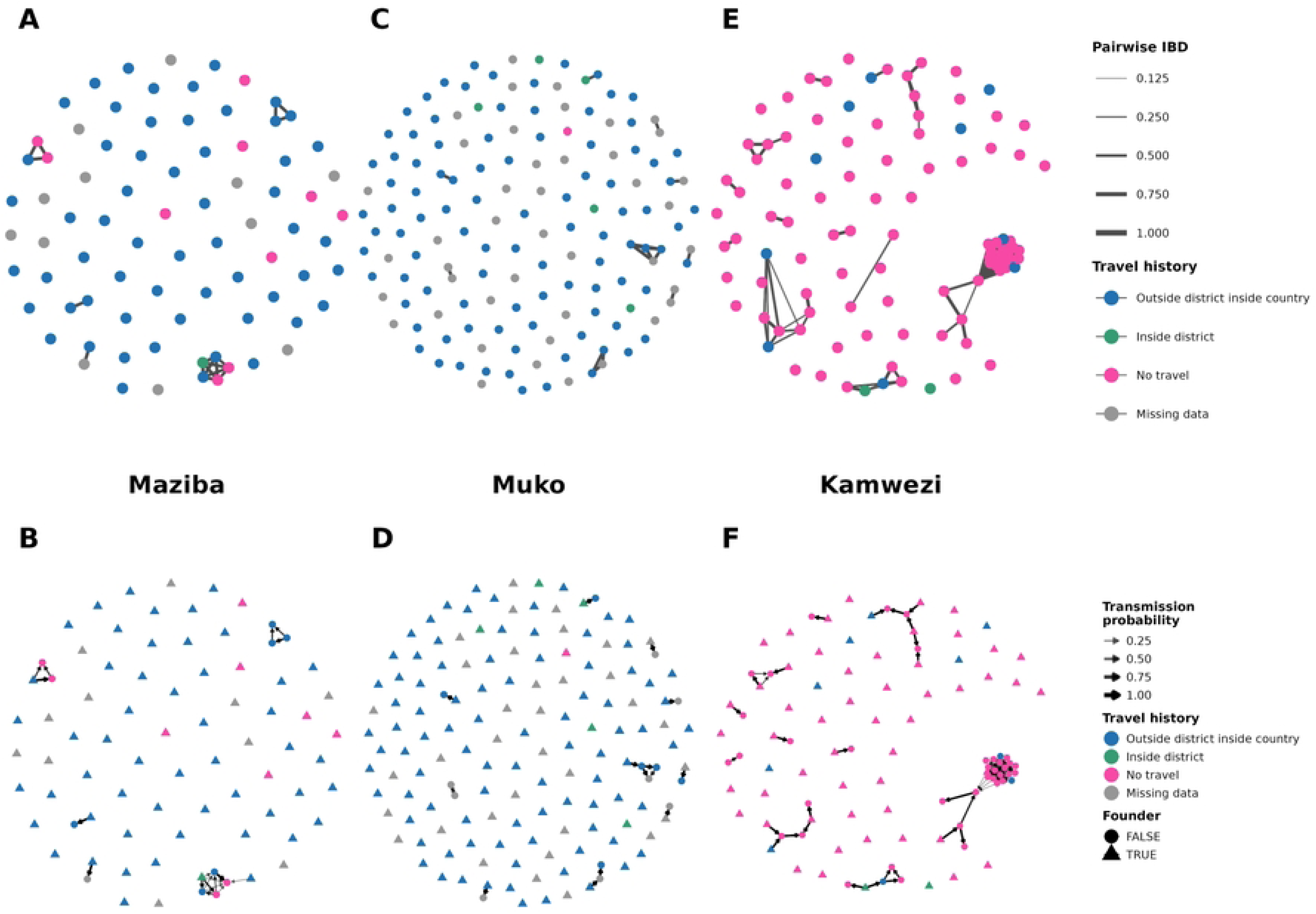
Genetic relatedness networks (Dcifer) and corresponding transmission networks for Maziba, Muko, and Kamwezi. Panels A, C, and E display pairwise parasite relatedness estimated using Dcifer. Each node represents an individual Plasmodium falciparum infection, and edges connect sample pairs with estimated relatedness (r) > 0.125. Edge thickness and color intensity are scaled to the magnitude of r, such that more strongly related infections appear as darker, thicker connections. Panels B, D, and F show model-based transmission networks for the same sites. Edges represent inferred transmission links, with edge width and transparency scaled continuously according to the posterior probability of direct transmission (0–1). Node color indicates reported travel history, and node shape distinguishes infections inferred to be founders (triangles) from non-founders (circles).

Accordingly, most infections in the very low transmission sites had no observed parents, (defined as founder infections in **Fig 2**), which is most consistent with importation of these cases. In Maziba, the model-estimated proportion of infections that were imported was 0.87 (95% credible interval (CrI): 0.80-0.94); in Muko, the estimated proportion imported was 0.91 (95% CrI: 0.86–0.95). These estimates likely represent an upper bound, as infections with unsampled parents, *e.g.*, those descended from imported infections which were asymptomatic, may be incorrectly classified as imported. Although travel was very common in Maziba and Muko, imported infections were significantly more likely to be associated with recent overnight travel compared to infections not classified as imported (OR = 6.8, 95% confidence interval (CI) 1.3-31.9). In addition, among infections with known travel status, all the source infections seeding transmission were individuals who reported recent travel; in two clusters in Maziba, people who reported no travel were infected by those who traveled (**Fig 2, panels B and D**). Therefore, genomic data from malaria cases in the low transmission sites shows a high proportion of imported infections and few, short transmission chains suggesting limited, sporadic local transmission, consistent with the high rates of travel reported at these sites.

In contrast, Kamwezi demonstrated higher levels of clustering consistent with sustained local transmission, including one large cluster of 30 related samples and several smaller clusters (**Fig 2E**). At this site, 30% of the samples belonged to at least one cluster. Due to the shorter sampling timeframe, and the fact that we did not attempt to capture all cases, it is likely the network analysis for Kamwezi represents an underestimate of the extent of local transmission. For this reason, the proportion of imported cases was not estimated from the transmission model. In Kamwezi, we observed a modest increase in the odds of being a source infection among those who reported travel, but differences were not statistically significant (OR = 1.9, 95% CI 0.3-9.1, **Fig 2F**). Genomic data from malaria cases in Kamwezi, characterized by higher relatedness and multiple inferred transmission chains, suggested substantially more local transmission than was evident at the lower-transmission sites, consistent with epidemiologic data indicating that few cases reported recent travel.

Finally, at Maziba and Muko, where samples were collected from malaria cases over 18 months, the mean R_e_ of each infection was estimated. Twenty infections had an estimated Rₑ > 0.8, indicating evidence of onward transmission. These infections were temporally clustered in both sites, occurring primarily from September 2023-January 2024, with a peak in November 2023 and again in April–June 2024, periods that aligned with elevated site-level incidence. Monthly incidence was positively associated with the odds of onward transmission (OR = 1.15; 95% CI 1.00–1.34), although this trend did not reach statistical significance. Though the small number of infections with evidence for onward transmission limits statistical power, the seasonal trend suggests that the data and model captured the underlying transmission dynamics expected during periods of higher incidence.

### Drug Resistance Markers

Genetic polymorphisms associated with artemisinin partial resistance (ART-R) have recently emerged and spread across Uganda. Analysis of *Pf*K13 polymorphisms showed that the P441L and R561H were the most prevalent mutations and were more common in Kamwezi than in Maziba and Muko (R561H significant for both comparisons [p<= 0.001], P441L significant only for Maziba [p = 0.03], **Table 3**). Other *Pf*K13 polymorphisms previously reported in Uganda, A675V, C469F, and C469Y, were present at lower frequencies across sites, while A578S and R539T were rare or absent (**Table S1** and **Fig S4**). A small number of polyclonal infections (n = 5) harbored more than one PfK13 mutation. Four samples carried two *PfK13* mutations, most commonly R561H co-occurring with C469F (n = 3), and one instance of A578S with P441L (n = 1). In addition, in one polyclonal infection three *PfK13* mutations (A578S, A675V, and P441L) were detected. To further assess how these variants were distributed within transmission networks in Kamwezi, we examined inferred transmission clusters with respect to *Pf*K13 mutations (**Fig 3**). The largest transmission cluster in Kamwezi was dominated by parasites carrying the P441L mutation (Panel A), whereas the R561H mutation (Panel B) occurred at a lower frequency in smaller clusters. Mixed *Pf*K13 genotypes observed within a few clusters were consistent with polyclonal infections (**Fig 3**).

**Table 3.**
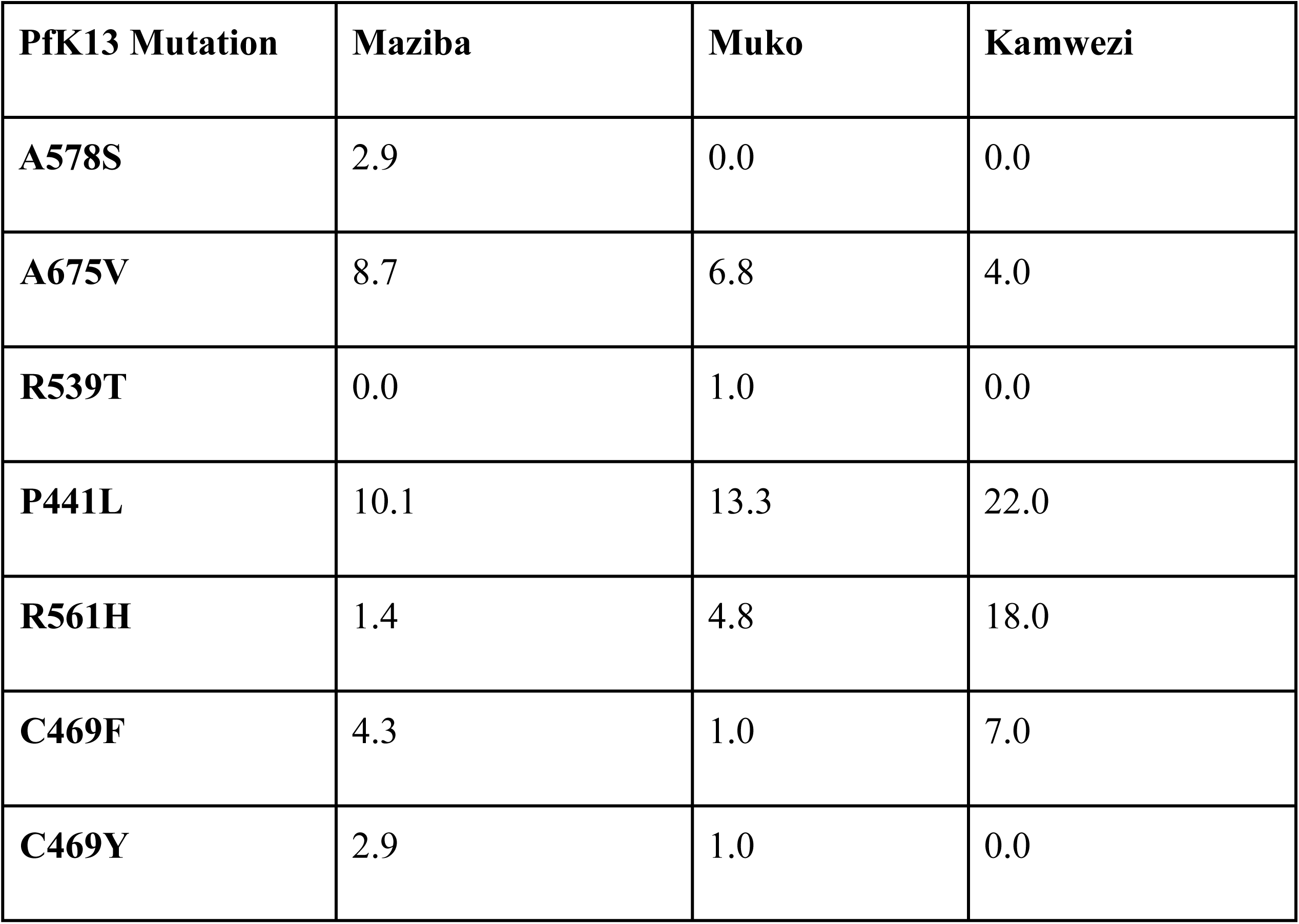
Prevalence of *PfK1*3 polymorphisms associated with ART-R.

**Fig 3.**
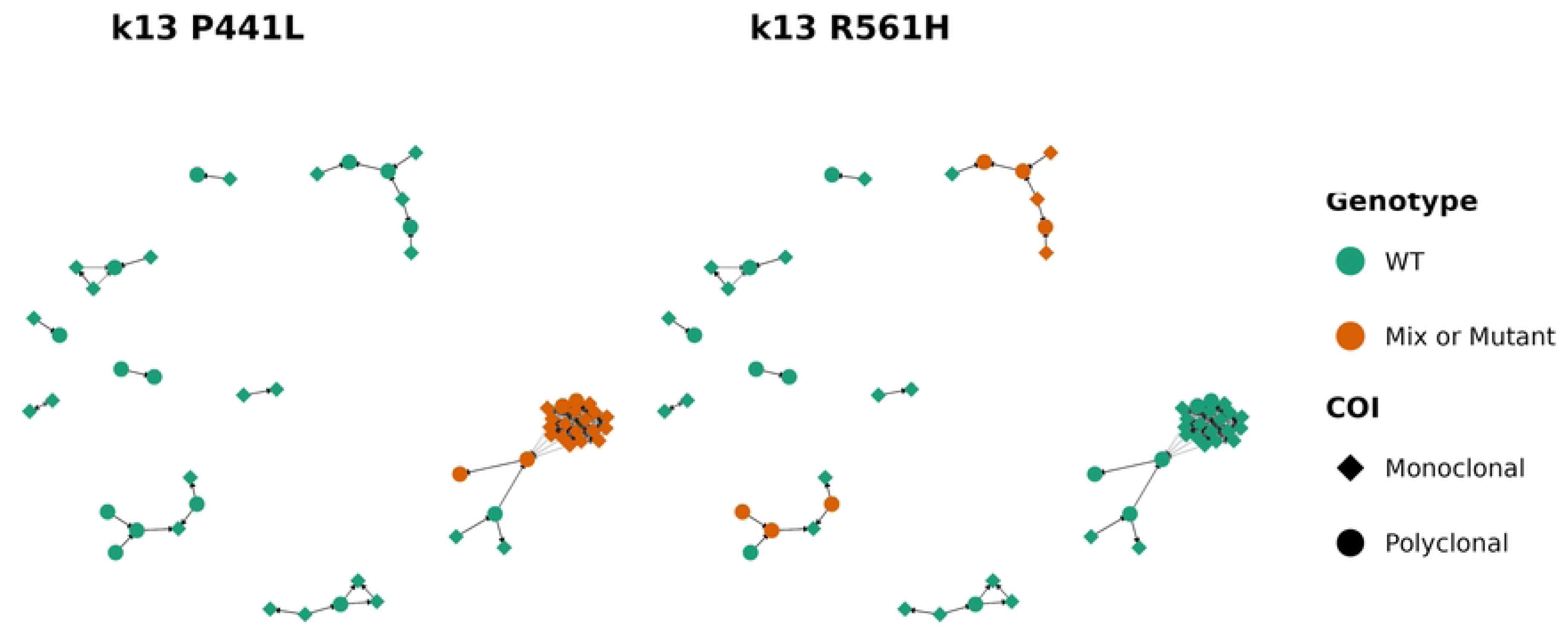
Transmission networks of P.falciparum in Kamwezi annotated for the observed PfK13 polymorphisms associated with ART-R. (A) Transmission network colored by K13 P441L genotype. (B) Transmission network colored by K13 R561H genotype. Nodes represent individual infections, with color indicating wild-type (green) versus mixed or mutant genotype (orange). Node shapes distinguish monoclonal (diamonds) from polyclonal (circles) infections. Arrows (directed edges) indicate inferred transmission direction based on posterior estimates from the transmission model.

We also profiled polymorphisms in other drug resistance genes, including *Pf*CRT*, Pf*DHFR*, Pf*DHPS, and *Pf*MDR1. As expected, markers associated with antifolate resistance (N51I, S108N, and K540E) were nearly fixed across all the three sites (**Table S1** and **Fig S4**). By contrast, other antifolate-associated alleles such as C59R and I164L exhibited more variation. Variants in *Pf*CRT and *Pf*MDR1 displayed only modest site-to-site fluctuations in allele frequencies without clear geographic structure. Notably, the *Pf*CRT K76T chloroquine-resistance allele was detected at low frequency in Muko and was absent in Maziba and Kamwezi. A full summary of all drug resistance-associated polymorphisms is provided in **Table S1** and visualized in **Fig S4**.

## Discussion

Integrated epidemiological and genomic analyses revealed distinct malaria transmission patterns across three health facilities in southwestern Uganda. At the very low transmission sites of Maziba and Muko, epidemiologic data - in particular, very high reported rates of travel in the 30 days prior to presentation with malaria - suggested importation. Consistent with this, infections in the low transmission sites had higher within-host diversity compared to infections in Kamwezi, likely reflecting acquisition in higher-transmission regions, where polyclonal infections are common. Further evidence of importation at Maziba and Muko was supported by transmission network analysis, which indicated that most cases were genetically unrelated and associated with travel, with evidence of minimal local onward transmission. In contrast, the epidemiologic and genomic data from Kamwezi were consistent with ongoing local transmission, with a smaller contribution from importation. Reported travel rates were much lower in Kamwezi, and infections exhibited lower genetic diversity but increased pairwise relatedness and well-defined transmission clusters, features indicative of localized spread. The younger age distribution of cases in Kamwezi further supports this interpretation, suggesting transmission is occurring within households and/or the community rather than being driven primarily by travel-related exposures.

Reported travel in the last 30 days among malaria cases enrolled at Maziba and Muko was strikingly high, surpassing rates observed in prior studies from Uganda that associated recent travel with malaria risk in low-endemic areas [38–40]. Similar patterns have been observed elsewhere in East Africa, including Ethiopia and Zanzibar, where importation continues to challenge elimination efforts [41,42], as well as in Southern Africa, where both internal and cross-border travel reintroduce parasites into areas approaching elimination [43]. Notably, the proportion of cases reporting recent travel at each site corresponded well with the estimated proportion of imported infections from the transmission network model, which classified cases as imported independent of travel data, using only dates of malaria diagnosis and genetic data. This highlights the usefulness of travel data as a proxy for importation and supports incorporating travel history into Uganda’s routine malaria case reporting in low-transmission areas. Genomic data added to the usefulness of the travel data by demonstrating very little local transmission at Maziba and Muko, with most imported infections leading to no other detected infections, and at most 2 or 3 generations of local transmission observed in any cluster. All of the source infections seeding the small transmission clusters in these sites were individuals who reported travel.

Other studies of low transmission areas have also shown a significant contribution of importation using genomic relatedness data. In Zanzibar, genomic evidence has shown that importation from mainland Tanzania is a significant contributor to malaria burden [44]. Likewise, a study of the parasite population from Bioko Island showed higher parasite diversity than expected given local transmission intensity and parasite relatedness between parasites on Bioko Island and mainland Africa suggestive of importation [45]. Similar findings have been reported in southern Mozambique, where genomic relatedness patterns combined with mobility data indicated that a substantial proportion of infections were likely imported from higher-transmission regions within Mozambique or neighboring countries [46]. Parasite populations in these settings showed high genetic diversity and limited clustering, consistent with repeated introduction of genetically distinct parasite strains rather than sustained local transmission. More recently, a study conducted on Pemba Island, Zanzibar from 2020–2022 found that 68% of reported malaria cases were classified as imported, based on travel history and epidemiological data [47]. Our findings in Maziba and Muko align with these patterns: both sites exhibited high parasite diversity, low within-site genetic relatedness, and only a few, short transmission chains, supporting the interpretation that importation plays a major role in sustaining transmission at these sites.

In contrast, patterns in Kamwezi indicated a higher contribution of local transmission to malaria burden. Compared with Maziba and Muko, a higher proportion of infections in Kamwezi were genetically related to one another, and transmission network analysis showed multiple large connected clusters. Incidence data from Kamwezi over the study period was consistent with epidemic, unstable malaria transmission, which likely reflected local ecological factors – such as more competent vector populations or microclimatic conditions favoring transmission, that sustain local outbreaks during specific periods of time even in a generally low-transmission region. A recent study in Kamwezi found that the association between travel to higher transmission areas and malaria risk was strongest during periods of low incidence [48]. This is consistent with our findings of predominantly local rather than imported transmission, as sample collection occurred during a period of relatively high local transmission. Evidence from other low transmission settings also supports the co-existence of both local transmission and importation. In Zanzibar, genomic analyses identified clear local transmission clusters despite substantial importation from mainland Tanzania, and similar patterns have been observed on Bioko Island, where locally related clusters were detected alongside signatures of imported infections [44,45]. Together, these findings indicate that during our sampling period, the contribution of local transmission was greater in Kamwezi than in Maziba and Muko. Given Kamwezi’s more variable, epidemic-prone transmission dynamics, malaria control efforts should include standard interventions such as vector control targeting local transmission, which will remain valuable during periods of elevated transmission intensity, along with importation-focused measures during periods of low transmission.

Differences in the distribution of PfK13 mutations associated with ART-R across the three sites illustrate how local transmission dynamics and regional connectivity may shape the spread of resistance. Although both P441L and R561H were detected at low levels in Maziba and Muko, their prevalence was substantially higher in Kamwezi. This contrast is consistent with site-level differences in importation. In Kamwezi, where there was more evidence of local transmission, resistance allele prevalence more closely reflected the previously published regional prevalence of these mutations (Conrad NEJM). Conrad et al., 2023 provide important national context for these patterns, showing that R561H and P441L have risen primarily in southwestern Uganda, with R561H increasing to 16–23% in Rukiga District and P441L reaching 12–23% at multiple western sites. In contrast, these mutations remain sporadic in northern and eastern Uganda, where other K13 variants dominate [49]. The elevated prevalence of P441L and R561H in Kamwezi is therefore consistent with its location within this southwestern hotspot and its proximity to Rwanda, where R561H first emerged before spreading into Uganda [49,50]. The P441L mutation, currently classified as a candidate resistance-associated variant, has also been increasing in frequency across multiple African settings [50–52], underscoring the need for continued functional and epidemiologic investigation to clarify its clinical relevance. These findings highlight the need to interpret drug-resistance patterns through the dual lens of local transmission dynamics and regional connectivity, and they emphasize the importance of targeted genomic surveillance to track the emergence and spread of resistance mutations.

Our study demonstrates the power of combining genomic and epidemiologic data to understand malaria transmission dynamics in low transmission regions. Most prior studies using genomic data to investigate the contribution of importation relied only on measures of pairwise relatedness between parasites or infections to identify potential evidence of local transmission.

Our study extended this methodology by using Plasmotrack, a Bayesian framework explicitly based on the biology of malaria transmission that infers directed edges between infections, identifying parent–offspring relationships and providing a statistical framework for estimating infection-level probability of importation and effective reproduction number. In addition, while travel history can indicate whether infections may have been imported, it cannot establish whether those infections generated onward transmission. Plasmotrack enabled us to address this gap by showing that while nearly all local transmission in Maziba and Muko was initiated by imported cases, these transmission clusters were small and unsustained. Furthermore, while reported travel history in our sites was consistent with the proportion of imported infections estimated by Plasmotrack, other studies have shown that genomic data can reveal connectivity not captured through travel surveys alone [12]. Transmission network analysis methods like Plasmotrack are, therefore, likely to be particularly valuable in these settings.

This study had some limitations. First, data collection was restricted to symptomatic malaria cases presenting to health facilities, without parallel community-based sampling. Thus, it did not capture asymptomatic infections or determine whether the high reported travel rates observed among malaria cases in Maziba and Muko reflected broader population-level mobility patterns. Second, the sampling periods differed across sites: Kamwezi was partially sampled over a two-month period, whereas Maziba and Muko were sampled continuously over a longer timeframe. These differences may have influenced comparisons of transmission dynamics across sites. Ongoing work is focused on more intensive sample collection and genotyping from malaria cases at Kamwezi over a longer time frame and expanding community-based sampling to address these gaps. Finally, even at the low transmission sites where we aimed to capture all malaria cases, we captured only approximately 60% of cases. For any transmission network analysis, deeper sampling will improve our ability to recapitulate transmission networks and result in more precise estimates of importation and reproduction number.

### Implications for malaria control and future directions

Uganda has recently finalized an ambitious National Malaria Elimination Strategic Plan (2026–2030) [53], which emphasizes sub-national stratification and tailored intervention packages in line with WHO’s “no one-size-fits-all” principle [54]. Within this framework, distinguishing between locally acquired and imported malaria infections is essential, particularly in low-transmission settings targeted for elimination. Our findings demonstrate that integrating epidemiologic surveillance, travel history, and parasite genomic data provides critical insights into malaria transmission dynamics and can inform more effective, context-specific, and evidence-based malaria control strategies in districts prioritized for elimination under the new national strategy.

These data can be directly leveraged to inform tailored intervention strategies. In sites like Muko and Maziba, where importation dominates, control efforts may need to focus on mobile populations. Potential strategies include targeted “test-and-treat” approaches for returning travelers, provision of chemoprophylaxis for individuals traveling to higher-transmission areas, and strengthened health education around personal protective measures during travel. In Kamwezi, by contrast, where substantial local transmission was observed alongside importation, a combined approach may be warranted, maintaining standard control interventions to decrease local transmission during high transmission periods, augmented by targeted measures to reduce imported infections during periods of lower transmission.

Notably, travel history collected through routine case management closely aligned with genomic estimates of importation across sites. This concordance highlights the value of systematically collecting travel data within routine health surveillance systems. The Uganda National Malaria Elimination Division may therefore consider incorporating standardized travel history questions into health facility reporting in low-endemic districts to better identify importation-driven transmission. Finally, building capacity for real-time genomic monitoring and embedding these data within national and regional surveillance frameworks will be essential to support timely public health responses, particularly in border and low-transmission districts where parasite movement threatens to undermine elimination gains. Sustaining progress towards malaria elimination in southwestern Uganda will require maintaining core interventions while leveraging genomic insights to anticipate and respond to evolving transmission and drug-resistance threats.

## Disclaimer

The funders had no involvement in the design of the study, data collection or analysis, decision to publish, or preparation of the manuscript.

## Data availability

All data underlying the findings of this study will be deposited in the NCBI public repository (Sequence Read Archive, SRA) and will be made freely available at the time of publication. The accession numbers will be provided upon acceptance.

## Acknowledgments

We thank all study participants for their invaluable contributions, as well as the dedicated field teams whose commitment ensured the successful implementation of this work. We are also grateful to the IMMRSE-U and ZUMBA study teams for their collaboration and support, and to colleagues at IDRC and UCSF for their mentorship and guidance throughout the study.

